# Overlapping transmission of group A and C/G *Streptococcus* facilitates inter-species mobile genetic element exchange

**DOI:** 10.1101/2023.08.17.23294027

**Authors:** Ouli Xie, Cameron Zachreson, Gerry Tonkin-Hill, David J Price, Jake A Lacey, Jacqueline M Morris, Malcolm I McDonald, Asha C Bowen, Philip M Giffard, Bart J Currie, Jonathan R Carapetis, Deborah C Holt, Stephen D Bentley, Mark R Davies, Steven YC Tong

**Author notes:** These authors contributed equally to the study (Mark R Davies, Steven YC Tong). Corresponding author: Prof Steven Tong The Peter Doherty Institute for Infection and Immunity 792 Elizabeth St, Melbourne, VIC, Australia, 3000.

## Abstract

*Streptococcus dysgalactiae* subspecies *equisimilis* (SDSE) and *Streptococcus pyogenes* share skin and throat niches with extensive genomic homology and horizontal gene transfer (HGT) possibly underlying shared disease phenotypes. It is unknown if cross-species transmission interaction occurs. We conducted a genomic analysis of a longitudinal household survey in remote Australian First Nations communities for patterns of cross-species transmission interaction and HGT. From 4,547 person-consultations, 294 SDSE and 315 *S. pyogenes* isolates were sequenced. SDSE and *S. pyogenes* transmission intersected extensively among households and the observed co-occurrence and transmission links were consistent with independent transmission without inter-species interference. At least one of three near-identical cross-species mobile genetic elements (MGEs) carrying antimicrobial resistance or streptodornase virulence genes was found in 55 (19%) SDSE and 23 (7%) *S. pyogenes* isolates. These findings demonstrate extensive co-circulation of both pathogens and HGT and support a need to integrate SDSE and *S. pyogenes* surveillance and control efforts.

## Introduction

*Streptococcus dysgalactiae* subspecies *equisimilis* (SDSE, commonly group C/G *Streptococcus*) is closely related to the better-known human pathogen, *Streptococcus pyogenes* (group A *Streptococcus*). SDSE shares much the same ecological niche on the human skin and throat as *S. pyogenes* and the two pathogens exhibit overlapping disease manifestations such as pharyngitis and invasive disease including necrotising fasciitis and streptococcal toxic shock syndrome^1^. In regions with a high burden of beta-haemolytic streptococcal disease and post-infectious sequelae, there has been evidence that superficial SDSE infection may trigger immune responses which cross-react with cardiac myosin^2, 3^. These findings raise the possibility that SDSE may contribute to immune priming and the burden of rheumatic heart disease in those regions^2^. In high income regions, emerging evidence has also described crude rates of invasive SDSE disease comparable to, and in some jurisdictions, greater than *S. pyogenes*^4–6^.

Whole genome comparisons of SDSE and *S. pyogenes* demonstrate extensive genomic homology including shared virulence factors such as the multi-functional surface M protein and evidence of horizontal gene transfer (HGT), frequently involving mobile genetic elements (MGEs)^7–9^. These similarities may contribute to shared disease phenotypes. Many *S. pyogenes* vaccine candidates are present in both species with evidence of cross-species homologous recombination^9^.

Despite extensive genomic homology, there is *in vitro* evidence of possible cross-species competition. Strains of the two pathogens possess shared quorum sensing genes such as the *sil* locus with evidence of cross-species signalling^10^. Furthermore, anti-microbial peptides or bacteriocins such as SpbN/SpbM and the SDSE-specific Dysgalacticin, are found in some strains of SDSE and *S. pyogenes* with cross-species activity^11, 12^.

SDSE and *S. pyogenes* transmit by common pathways including respiratory droplets^1^. Recently, we have shown that asymptomatic *S. pyogenes* throat carriage is an important reservoir of transmission in high-endemic settings^13^. Transmission pathways of SDSE have not previously been described. Further, it is uncertain if in real-world studies transmission of one species competes with the other. In communities endemic for *S. pyogenes* infection with high rates of skin infection, rheumatic heart disease and invasive disease, the current focus is largely on *S. pyogenes* control through skin sore and scabies control programs, and vaccine development. Understanding the transmission interactions of SDSE and *S. pyogenes* and anticipating the potential impact of disease control measures on cross-species behaviour is important to inform the design of surveillance programs and infection control efforts.

In this study, we examine the transmission of SDSE at a whole genome sequence (WGS) resolution using isolates collected in a household-based surveillance study over two years in two remote communities in the Northern Territory of Australia^14^. These transmission networks were compared to that of co-collected *S. pyogenes* isolates to assess for inter-species transmission interactions, and in the setting of co-circulation, their genomes were systemically examined for evidence of cross-species HGT of MGEs carrying key virulence and antimicrobial resistance genes.

## Results

### Sampling and clinical epidemiology

Two remote Aboriginal communities in the Northern Territory of Australia were prospectively followed for a two-year period between 2003–2005^14, 15^. Observations for one community (community 3) commenced in June 2004 as it replaced an initial community (community 2) with low recruitment. Communities 1 and 3 were included in this study.

Households (18 in community 1 and 20 in community 3) were visited approximately monthly allowing for access affected by weather and cultural events (Supplementary Figure 1). At each visit, throat swabs were taken regardless of symptoms and skin swabs were taken from impetigo lesions. From a total of 4,547 person-consultations during 486 household-visits, 1,087 individuals (547 from community 1 and 540 from community 3) were sampled from which 330 SDSE isolates (252 community 1 and 78 community 3) were recovered. Of the 330 isolates, 8 were from skin and 322 from throat swabs of which only one case reported a sore throat. *S. pyogenes* was recovered on 327 occasions (218 community 1 and 109 community 3) with 208 isolated from throat swabs and 119 from impetigo lesions. Detailed descriptions of the epidemiology of cases were described previously^14, 15^.

There was a high rate of individual mobility in and out of households with a median of 28 people (range 6–57) enrolled per household over the study period. Each individual was observed at a median of 3 visits (range 1–19, intermittently sampled); and as such, duration of carriage in individuals could not be determined. Households were positive (i.e., at least one individual positive) for SDSE for a median of 56 days and then re-acquired SDSE a median of 37 days later.

### Whole genome sequencing reveals detailed transmission clusters

From the 330 SDSE isolates, 294 (89%) were recovered for WGS. Using traditional epidemiological markers, *emm* type and multilocus sequence type (MLST), these isolates represented 19 *emm* types (23 *emm* subtypes), 21 MLSTs, and 26 *emm*-MLST combinations (Supplementary Table 1). Of these, 8/26 (31%) *emm*-MLST groups were found across both communities. Sequencing and analysis of 315/327 (96%) *S. pyogenes* isolates recovered from communities 1 and 3 for WGS were reported previously^13^.

To determine a WGS threshold for clustering of strains, we examined genomic variation of isolates of the same strain found longitudinally on multiple occasions from the same individual. Intra-host variation was used to predict longitudinal diversity of strains forming transmission chain as well as technical variations in single nucleotide polymorphism (SNP) calling. SDSE was found in 58 individuals on more than one occasion including three who were positive on five occasions, four on four occasions, 15 on three occasions and 36 on two occasions. Using *emm* and MLST as markers, 36 individuals had the same strain on more than one occasion including six individuals with the same isolate on three occasions, one individual on four occasions, and one on five occasions (Supplementary Figure 2). Pairwise SNP distances were calculated between these isolates and a threshold of <8 SNPs was determined for WGS transmission clustering (Supplementary Figure 3).

Phylogenetic reconstruction supported 18 distinct SDSE lineages/global genomic sequence clusters^9^ present across both communities (Figure 1). High resolution genomic transmission clusters based on single linkage clustering at a SNP threshold of <8 and >99% shared gene content, revealed much finer detail than the traditional epidemiological markers (Figure 2). A total of 37 SDSE transmission clusters representing 237 (81%) isolates were inferred with an additional 57 singleton isolates (Supplementary Table 1). Transmission clusters were supported by core SNP phylogenies and presence-absence of virulence and/or antimicrobial resistance genes (Supplementary Figure 4a-c) with significant diversity within *emm* types (Figure 2) and evidence of mobile genetic element (MGE) gain/loss events carrying antimicrobial resistance and/or virulence factor genes among closely related isolates.

**Figure 1.**
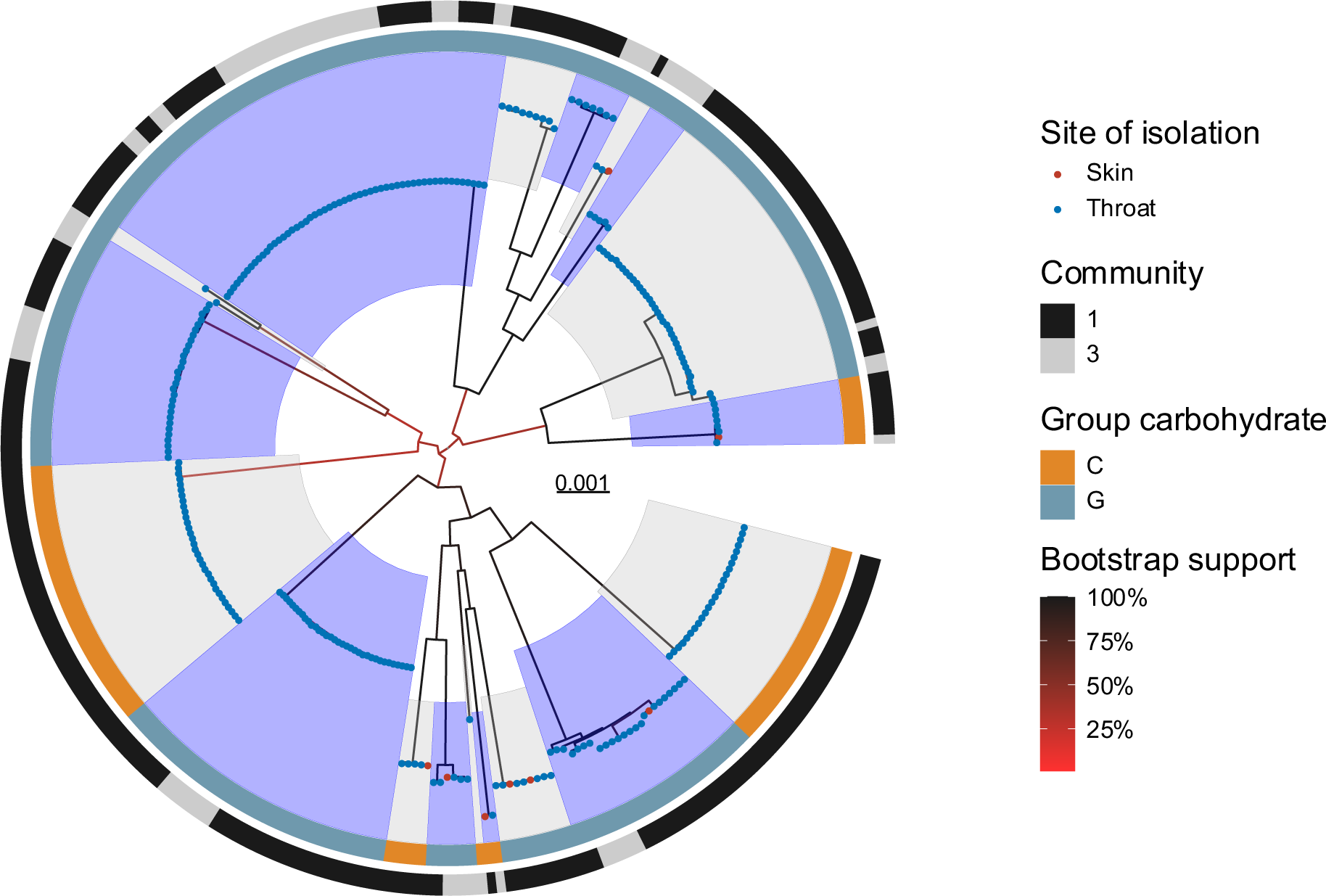
Maximum-likelihood phylogeny of 294 *Streptococcus dysgalactiae* subsp. *equisimilis* (SDSE) isolates. Sequences were aligned against SDSE reference genome GGS_124 (NC_012891.1) with mobile genetic element regions masked. Distinct genomic sequence clusters determined by PopPUNK^25^ as previously defined by a global SDSE dataset^9^, are denoted by alternating blue and grey highlights from internal nodes. Site of isolation is coloured by blue (throat) and red (skin) tips. The inner ring denotes the Lancefield group carbohydrate and the outer ring the community of isolation. Bootstrap supports are shown as branch colour gradients and were calculated using the ultrafast bootstrap approximation demonstrating some uncertainty in deep branches of the phylogeny^30^.

**Figure 2.**
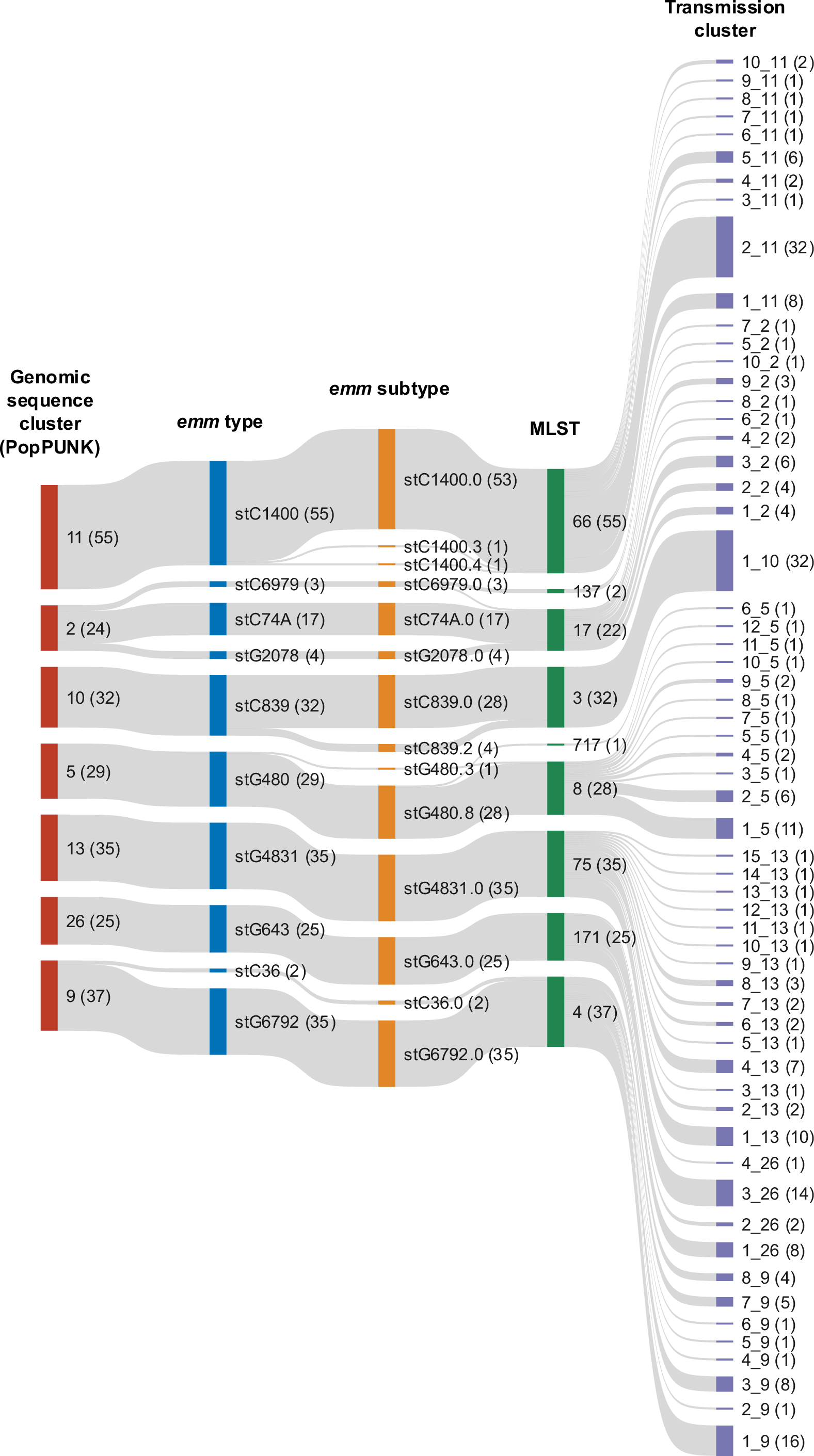
Alluvial plot of the relationship between the largest 7/18 *Streptococcus dysgalactiae* subspecies *equisimilis* (SDSE) genomic sequence clusters (representing 237/294 isolates) as determined by PopPUNK against *emm* type, *emm* subtype, multilocus sequence type (MLST) and transmission clusters determined using single linkage clustering at a SNP threshold of <8 and >99% shared gene content. The number of isolates in each category is denoted within brackets. From 10 *emm* types (14 *emm* subtypes) and 9 MLSTs shown, 60 transmission clusters were determined. Traditional markers such as *emm* subtype in some cases over-split SDSE clusters as demonstrated by *stC*839.0 and *stC*839.2 which differ by only one SNP within their hypervariable *emm* region and otherwise fall within the same transmission cluster.

The two largest transmission clusters consisted of 32 isolates each and clusters with four or more isolates made up a total of 204 isolates (69%). Transmission clusters were present across a mean of 3 households (range 1–16). Despite the finding of eight *emm*-MLST groups across both communities, the WGS analysis indicated that only a single transmission cluster spanned both communities. The upper limit of the pairwise SNP distance between isolates of the same transmission cluster was 16 SNPs (median 4) compared to 791 SNPs (median 20) within the same MLST, 5491 SNPs (median 25) within the same *emm* type, 638 SNPs (median 19) within the same *emm*-MLST combination, and 1505 SNPs (median 21) within the same genomic sequence cluster (Supplementary Figure 5), highlighting the limitations of other markers in determining recent transmission clusters.

There was no clear pattern of *emm* type replacement of SDSE isolates over time in the two communities in contrast to sequential replacement of *S. pyogenes emm* types as reported previously^13, 14^. Consistent with this finding, SDSE transmission clusters persisted for longer in the two communities (median of 349 days, 95% CI 189-440 days) compared to *S. pyogenes* (median of 241 days, 95% CI 181-259 days, log-rank p = 0.009) (Supplementary Figure 6).

### Network analysis supports independent transmission dynamics for SDSE and S. pyogenes

SDSE transmission between households within each community was modelled by inferring links between isolates of the same transmission cluster detected at successive community visits (transmission window 12-44 days), including intra-household transmission events. Individuals were grouped by household which formed the nodes of the transmission work. Analysis of the transmission network revealed 123 SDSE putative transmission edges (events) in community 1 and 14 edges in community 3, which had a shorter duration of sampling and fewer isolates detected (Table 1). All but one transmission edge was attributed to isolates from throat swabs for SDSE in contrast to 50/173 (29%) edges attributed to a predicted skin source for *S. pyogenes*.

**Table 1.** Number of inferred unweighted transmission edges between households at successive community visits in two remote communities.

| *Streptococcus dysgalactiae* subspecies *equisimilis* |  |  |  |  |  |
| --- | --- | --- | --- | --- | --- |
|  | Household |  | Community |  | Total |
|  | All isolates | Throat source | All isolates | Throat source |  |
| Community 1 | 22 (18%) | 22 (18%) | 101 (82%) | 100 (81%) | 123 |
| Community 3 | 3 (21%) | 3 (21%) | 11 (79%) | 11 (79%) | 14 |
| *Streptococcus pyogenes* |  |  |  |  |  |
|  | Household |  | Community |  | Total |
|  | All isolates | Throat source | All isolates | Throat source |  |
| Community 1 | 15 (11%) | 9 (7%) | 116 (89%) | 75 (57%) | 131 |
| Community 3 | 4 (10%) | 3 (7%) | 38 (90%) | 36 (86%) | 42 |

To test the hypothesis that transmission of SDSE or *S. pyogenes* may interfere with transmission of the other species, the overlap between inferred transmission networks of the two species was compared to a null model in which any cross-species interaction was removed.

“Transmission overlap” was defined as the proportion of inferred SDSE transmission edges that corresponded to an inferred transmission of *S. pyogenes*. An overlapping edge corresponded to transmission of both SDSE and *S. pyogenes* which occurred between the same pair of households within the same transmission window without distinguishing which household acted as source. To generate a null model of transmission overlap, household labels in the inferred SDSE transmission network were randomised while preserving the *S. pyogenes* network. This process preserves important structural features of the SDSE network including degree distribution, and any clustering of SDSE transmission between households, while removing any direct cross-species effects related to the transmission of *S. pyogenes*.

Overlaying the transmission networks of the two species found a highly interconnected network with 11 shared transmission edges – nine in community 1 and two in community 3 (Figure 3). The number of shared edges in each community was consistent with the distribution under the null model providing no evidence of inter-species transmission interference (one-sided p-value ≤ observed value for community 1 = 0.75, community 3 = 0.94) (Supplementary Figure 7a, c). Results were similar when restricting the analysis to isolates only from throat swabs (Supplementary Figure 7b, d). These results indicate no evidence of an interaction between the two species in their household transmission patterns.

**Figure 3.**
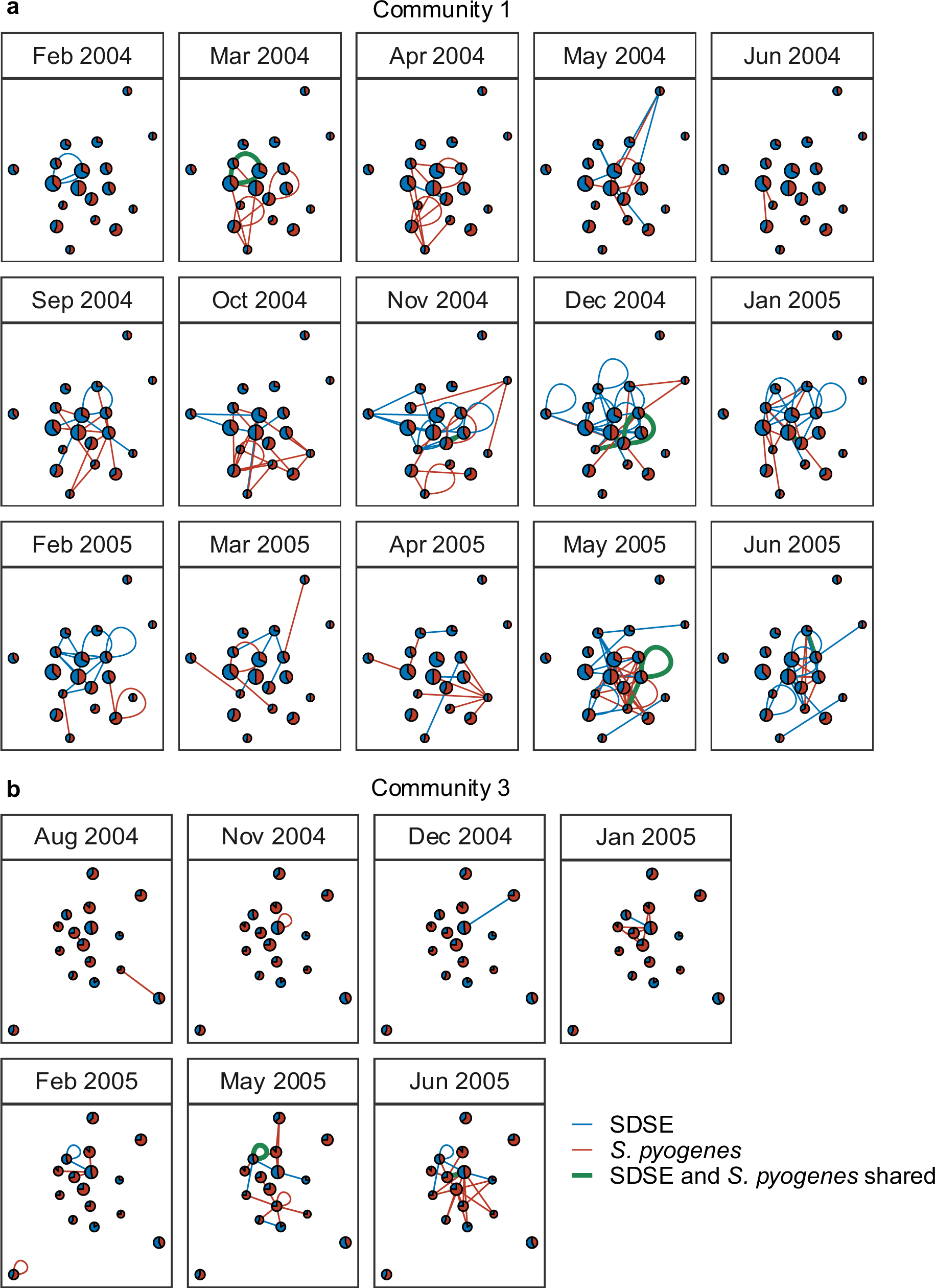
Transmission links between households across consecutive community visits in community 1 **a)**, and community 3 **b)**. Households are represented by nodes proportional in size to the number of participants enrolled at each household and coloured by the proportion of *S. pyogenes* (red) and *S. dysgalactiae* subsp. *dysgalactiae* (SDSE, blue) isolates detected in the household across the entire study period. Transmission links are represented by undirected and unweighted edges between households and coloured by species with shared edges highlighted in green. Loops correspond to predicted transmission edges between unique individuals within the same household. Only community visits where transmission edges were predicted are shown.

Although only 11/137 (8%) of total SDSE transmission edges were shared with *S. pyogenes*, the combined transmission networks demonstrated extensive crossover of the two organisms at the household level — SDSE and *S. pyogenes* co-occurred in the same household on 100/486 (21%) of household-visits (Figure 4). To infer a null model of co-occurrence of SDSE and *S. pyogenes* in households while removing cross-species transmission effects, SDSE and *S. pyogenes* positive swabs were randomised across all swabs at each community visit. To account for grouping of isolates within households, isolates from the same transmission cluster were collapsed to a single positive result during the same household-visit. The observed co-occurrence of SDSE and *S. pyogenes* within households was consistent with the model of independent inter-species transmission without evidence of interference (one-sided p-value ≤ observed value across both communities = 0.62). Results from a sensitivity analysis limited to isolates from throat swabs were consistent (Supplementary Figure 8a, b).

**Figure 4.**
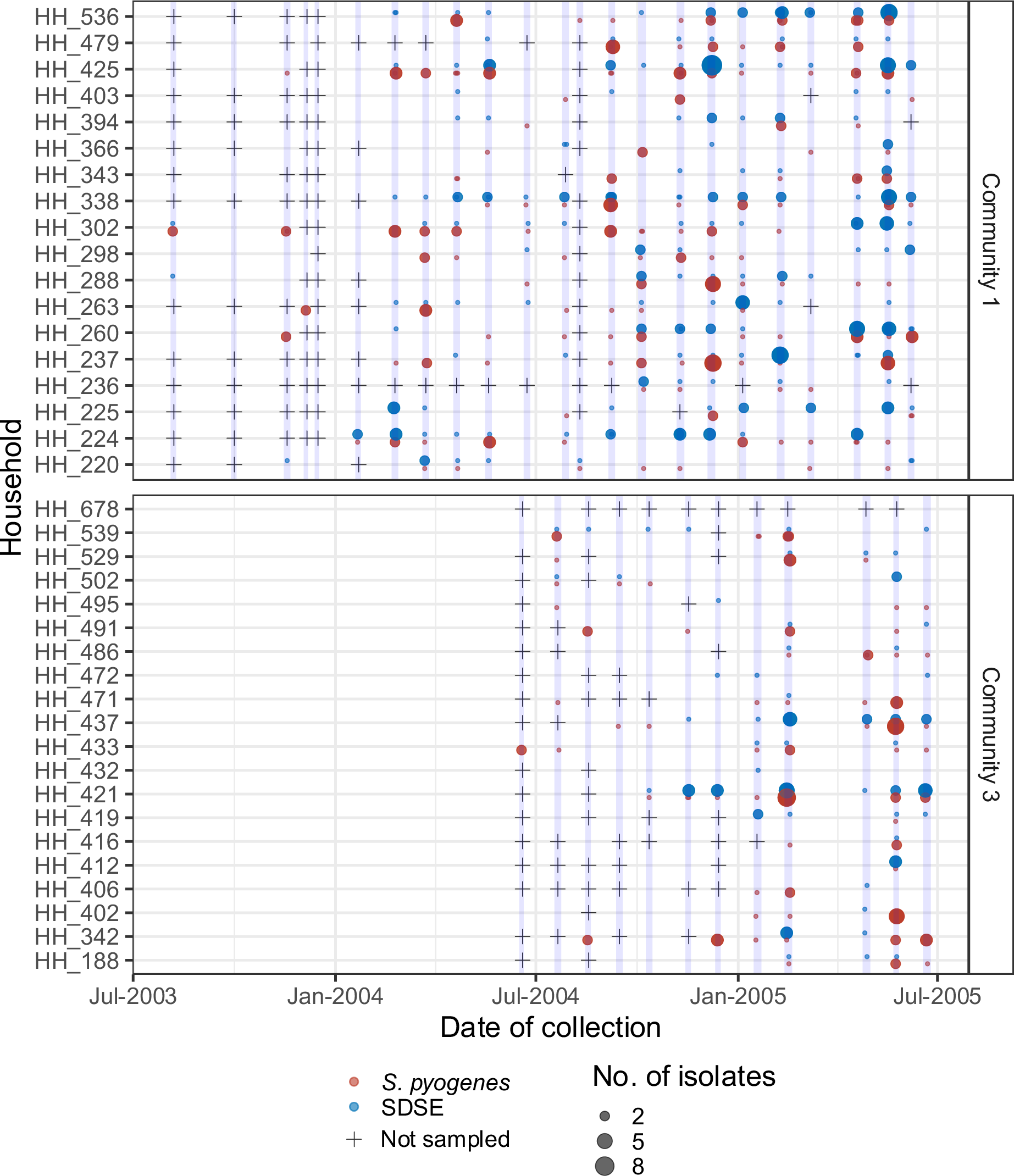
Co-occurrence of *Streptococcus dysgalactiae* subsp. *equisimilis* (SDSE) and *S. pyogenes* in households in communities 1 and 3 at each community visit (light blue highlights) during the study period. Detection of SDSE (blue) and *S. pyogenes* (red) are denoted by points with the size of each point proportional to the number of isolates. Community visits where a household was not sampled are denoted by crosses. SDSE and *S. pyogenes* co-occurred on 100/486 (21%) of household-visits.

### Co-occurrence of SDSE and S. pyogenes facilitates shared mobile genetic elements

We have previously demonstrated extensive genomic overlap between SDSE and *S. pyogenes* in the context of global genome databases^9^. In the setting of extensive household co-occurrence of the two species, we sought to find evidence of shared MGEs between the two species. Using a pangenome synteny-based approach, MGEs were systemically extracted from both SDSE and *S. pyogenes* isolates and examined for elements with >99% nucleotide identity across species^9, 16^.

Three near identical MGEs were found to be present in SDSE and *S. pyogenes* with variable presence across closely related isolates with as few as 0-11 core SNPs, suggestive of recent MGE gain/loss events within each of these strains (Figure 5a). A 53kbp prophage, ϕ1207.3^17^, carrying *mef(A)*/*msr(D)* macrolide efflux resistance genes was carried at a conserved cross-species genomic location (between SDEG_RS07105 and SDEG_RS07110 in reference genome GGS_124 NC_012891.1) and was present in 5 *S. pyogenes* and 31 SDSE isolates (Figure 5b). A second prophage, ϕMGAS5005.3 carrying the streptodornase gene *sda1*, previously described to be shared across species, was also found in a cross-species conserved insertion region^9^. An 18kbp integrative conjugative element (ICE)-like segment carrying the tetracycline resistance gene, *tet(M)*, was present in four *S. pyogenes* and eight SDSE isolates at three distinct insertion regions (Figure 5c, Supplementary Figure 9). At least one of these MGEs which carried antimicrobial resistance-associated genes or virulence-encoding genes, was found in 55 (19%) of SDSE and 23 (7%) of *S. pyogenes* isolates. SDSE isolates carrying these shared MGEs were found across both communities while *S. pyogenes* isolates carrying shared MGEs were restricted to single communities (community 1 for ϕ1207.3, community 3 for ϕMGAS5005.3 and the ICE-like element).

**Figure 5.**
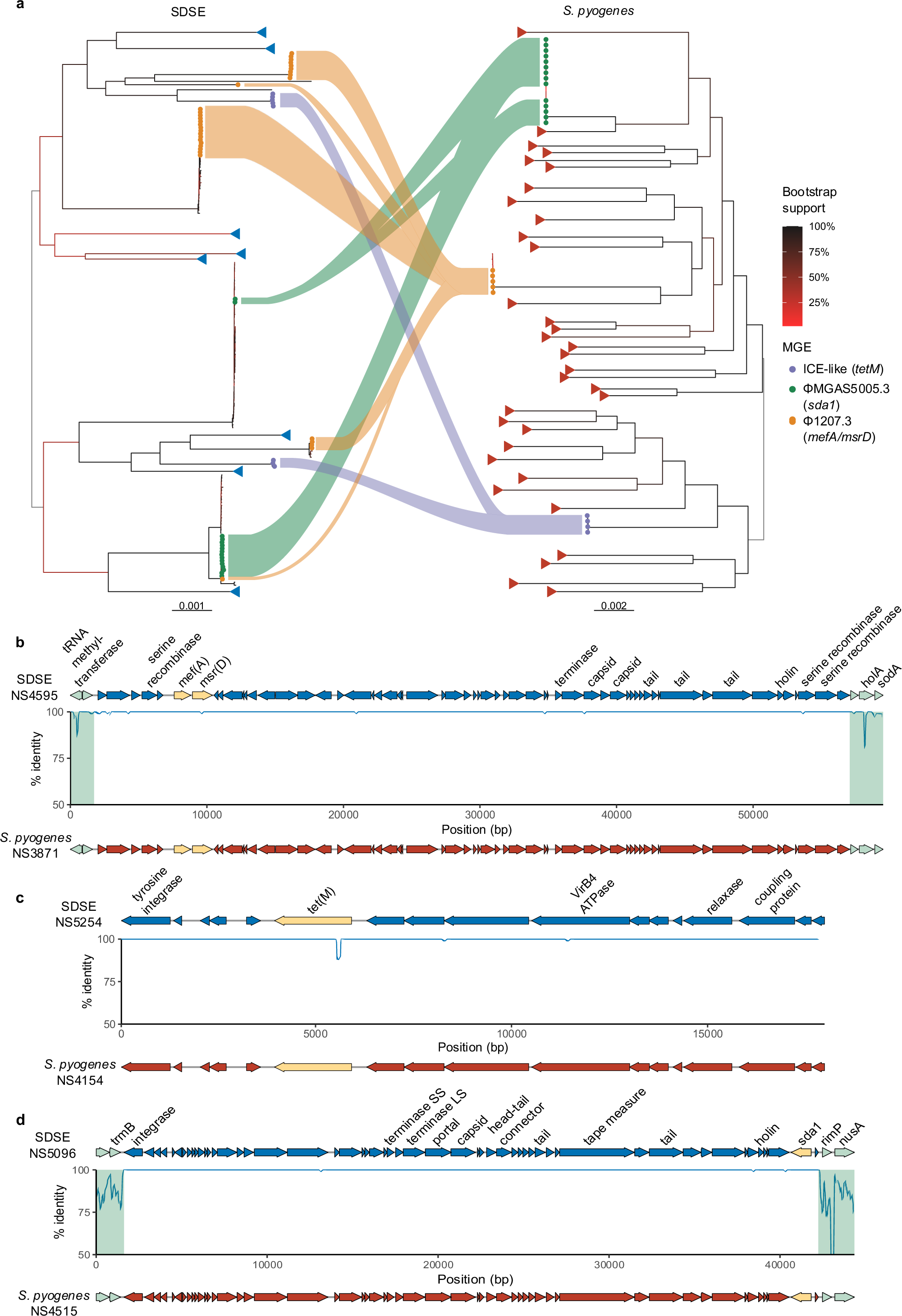
Shared mobile genetic elements (MGE) across *Streptococcus dysgalactiae* subsp. *equisimilis* (SDSE) and *Streptococcus pyogenes* isolates. **a)** Maximum likelihood trees of SDSE and *S. pyogenes* with isolates carrying three near-identical (>99% nucleotide identity) MGEs highlighted by tree tip points. Genomic sequence clusters without the three MGEs of interest are collapsed and denoted by blue (SDSE) and red (*S. pyogenes*) triangles at tree tips. Flows link corresponding shared MGEs across the species but do not imply directionality of transfer. Bootstrap supports are shown as branch colour gradients and were calculated using ultrafast bootstrap approximation^30^. **b)** A 54kbp prophage, ϕ1207.3, carrying macrolide efflux resistance genes *mef(A)* and *msr(D)* was present with >99.9% nucleotide identity across SDSE and *S. pyogenes*. A representative SDSE element from isolate NS4595 was aligned against a representative *S. pyogenes* sequence (NS3871) with percentage nucleotide identity calculated using Hamming distance and plotted in 100bp sliding windows. The element was present in a cross-species conserved insertion region with flanking core genes highlighted in green. ϕ1207.3 was present in 5/8 *emm*58.8, MLST 549 *S. pyogenes* isolates with as few as 11 single nucleotide polymorphisms (SNPs) between isolates with and without the prophage. The same element was present in 31 SDSE isolates across subsets of five different lineages suggestive of recent gain/loss events of ϕ1207.3 in both the SDSE and *S. pyogenes* populations. **c)** An 18kbp integrative conjugative element (ICE)-like MGE carrying the tetracycline resistance gene, *tet(M)*, was present with >99.9% nucleotide identity in 4 *S. pyogenes* and 8 SDSE isolates, including two distinct SDSE populations. The element was present at three different genomic insertion regions and thus flanking core genes are not shown. In the example shown, a 12bp in-frame deletion was present at the 5’ end of *tet(M)* in the SDSE element which was distant from the active ribosomal binding domain. **d) A** 41kbp prophage ϕMGAS5005.3 carrying the streptodornase gene *sda1*, was shared across species with >99.9% nucleotide identity at a cross-species conserved insertion region as has been described previously^9^. ϕMGAS5005.3 was present in 14/16 *S. pyogenes emm*1.0, MLST 28 isolates with a maximum of 3 SNPs between isolates and 16 SDSE isolates across subsets of two different lineages.

While directionality of MGE movement could not be inferred, including distinguishing between inter-species versus intra-species dissemination, the presence of near-identical elements at conserved insertion regions, suggests that overlapping transmission may facilitate shared MGEs from a common pool. The carriage of these MGEs across multiple distinct lineages suggests that these shared MGEs may lead to dissemination of antimicrobial resistance and virulence-associated genes.

## Discussion

Using WGS-level resolution, we were able to reconstruct SDSE household transmission networks and compare it to co-collected *S. pyogenes* isolates, demonstrating extensive co-circulation. Despite occupying similar niches on the skin and throat, we show that the two organisms transmit independently without evidence of interference at the household level. In the setting of extensive transmission cross-over in households, we find multiple MGEs present across both populations carrying antimicrobial resistance or virulence factor genes with evidence suggestive of recent gain/loss events. This analysis of a dataset of densely co-sampled SDSE and *S. pyogenes* isolates provides a level of transmission detail and examination of real-world inter-species transmission dynamics and horizontal gene transfer which to our knowledge, has not previously been described for beta-haemolytic streptococci.

SDSE is increasingly being recognised as an important cause of invasive human disease with recent studies suggesting incidence and mortality comparable to *S. pyogenes*^4–6^. While not traditionally considered as a cause of acute rheumatic fever/rheumatic heart disease (ARF/RHD), reports from northern Australia suggest that at least in high-incidence areas of ARF/RHD, SDSE throat carriage may have the potential to induce cardiac myosin cross-reactive antibodies mimicking that seen with *S. pyogenes*^2, 3^. Therefore, the finding of extensive throat transmission of SDSE, including persistence of transmission clusters longer than that of *S. pyogenes,* underscores a need to further understand its contribution to immune priming for ARF/RHD which in turn has important disease control implications.

Additionally, interactions between SDSE and *S. pyogenes* such as horizontal gene transfer and homologous recombination are key drivers in bacterial population dynamics, and may influence *S. pyogenes* and SDSE biology^9^. Notably, genes encoding antigens currently under investigation as *S. pyogenes* vaccine candidates are frequently also found in SDSE^9^. Our findings of extensive household co-occurrence may provide an opportunity for HGT which we demonstrate in the setting of shared MGEs. We show three near-identical MGEs were present across different lineages in SDSE and *S. pyogenes* including presence and absence in closely related isolates suggestive of recent gain/loss events. These MGEs carried antimicrobial resistance and virulence genes such as the macrolide efflux genes *mef(A)*/*msr(D)*, tetracycline resistance *tet(M)*, and the streptodornase gene *sda1*. While we cannot infer directionality of HGT of MGEs across species compared to intra-species dissemination or acquisition from an intermediary species, at least one of these MGEs was present in 55 (19%) of SDSE and 23 (7%) of *S. pyogenes* isolates. This underscores the importance of integrating SDSE with *S. pyogenes* surveillance as we seek to improve our understanding of transmission and disease pathogenesis of the two organisms and as efforts move towards a possible *S. pyogenes* vaccine which may introduce selection pressures across both organisms.

SDSE and *S. pyogenes* occupy similar ecological niches in the throat and on the skin with overlapping disease manifestations such as pharyngitis. Cross-species interaction and competition has been demonstrated such as the expression of bacteriocins which are able to inhibit the other species and cross-species quorum sensing involving the two-component regulator, *silAB* with its signalling peptide *silCR*^10–12^. However, the *sil* locus and characterised bacteriocins such as Dysgalacticin and SpbN/SpbM are variably present in SDSE and *S. pyogenes* and it is unclear if *in vitro* interactions translate to real-world transmission dynamics. Our data demonstrate that despite evidence of possible *in vitro* interference, SDSE and *S. pyogenes* appear to transmit independently with highly interconnected household transmission networks in a high burden setting.

SDSE was almost exclusively isolated from the throat in this study^14^. The mechanism behind the predilection for the throat for SDSE in comparison to the wider presence of *S. pyogenes* across throat and impetigo lesions is unclear. As described previously, the age of individuals included in this study with SDSE was not different to those with *S. pyogenes* with the highest rates in 5-14 year-olds and does not explain the throat predominance of SDSE^14, 15^. Despite the genomic similarities between SDSE and *S. pyogenes*, their virulence repertoires differ including carriage of the cysteine proteinase SpeB which is exclusively present in *S. pyogenes*. Experimental evidence suggests that SpeB activity may be important in establishing skin infection for *S. pyogenes*^18^. Cross-species genotype-phenotype associations could not be drawn from this study due to the near perfect separation between skin and throat sites for SDSE. However, sensitivity analyses restricting cross-species transmission analyses to throat isolates were concordant with the primary analysis without any evidence of cross-species interference. Despite evidence of independent transmission at a household level, with household co-occurrence of SDSE and *S. pyogenes* on 100/486 (21%) of household-visits, the frequency of presence of SDSE and *S. pyogenes* in the same swab is unclear. SDSE and *S. pyogenes* are both large colony, beta-haemolytic streptococci and are generally indistinguishable by colony morphology. Given only representative colonies were characterised in this study, the frequency of co-colonisation of SDSE and *S. pyogenes* in the same individual could not be estimated. In fact, this is a common limitation of carriage studies to date seeking to determine the prevalence of SDSE and *S. pyogenes* from throat swabs^19–21^. Given our findings of household-level transmission dynamics, future studies should consider methods such as WGS from plate sweeps or deep sequencing of swabs to determine co-occurrence in individuals. These methods have also previously been shown to improve resolution of intra-host diversity and reconstructing transmission and may offer greater insight into cross-species transmission dynamics^22^.

Our study has some limitations. This study was carried out in a remote and tropical setting in northern Australia in Aboriginal communities with a high burden of *S. pyogenes* disease including impetigo, ARF/RHD and invasive disease. Therefore, transmission dynamics and co-occurrence of the two organisms may differ in other settings. There was a high level of population mobility in and out of households in these communities and thus individual level transmission dynamics and duration of carriage could not be determined due to limited longitudinal sampling of most individuals. Additionally, while SDSE was only found from 8 impetigo/skin sore swabs, intact skin was not sampled. Therefore, it is unclear if SDSE on healthy skin may contribute to transmission.

In summary, this study demonstrates important transmission dynamics of SDSE and *S. pyogenes*. The two closely related pathogens frequently co-occur within households with interconnected transmission networks, but without evidence of inter-species interference across households. Transmission overlap and shared niches, particularly in the human throat, may facilitate interspecies gene flow including clinically important determinants such as antimicrobial resistance genes. These findings emphasise a need to further understand the interactions between these pathogens including in the context of ARF/RHD in high burden regions. The immunopathogenesis of ARF remains poorly understood despite many decades of research and the specific events antecedent to each episode of ARF are elusive with respect to the role of *S. pyogenes* in skin lesions and SDSE in the throat. As interventions targeting *S. pyogenes* take place, it is possible that SDSE may also be affected. That impact could potentially be a reduction in SDSE disease (e.g., by vaccines that may target common antigens) or conversely by SDSE filling an ecological niche if *S. pyogenes* infection or carriage is selectively targeted (e.g., in primary care interventions that expand the use of *S. pyogenes* rapid diagnostics for throat swabs). Incorporating research, surveillance and control efforts of SDSE with *S. pyogenes* will improve the understanding of both pathogens individually and cross-species interactions in relation to clinical disease burden, disease phenotypes, and future response to vaccine interventions.

## Methods

### Isolate collection and culture

Isolates were collected from a previously reported prospective surveillance study in three remote Aboriginal communities in remote Northern Territory, Australia, which were visited approximately monthly over a two-year period from August 2003 to June 2005^14^. Due to waning community support and logistical difficulties in community 2, it was replaced with another community in June 2004 (community 3). Only communities 1 and 3 were included in this study. At each visit, researchers collected throat swabs regardless of symptoms from participants and examined for skin sores both purulent and dry, which were also swabbed. Due to high population mobility, individuals were identified as part of households for analyses, including family groups residing in one or two adjacent houses.

Swabs were inoculated onto horse blood agar and selective media containing colistin and nalidixic acid and transported for culture at a central laboratory in Darwin, Australia. Plates were incubated at 37°C in 5% CO_2_ and examined after 24 and 48 hours. A single representative colony was selected for typing (Streptococcal Grouping Kit, Oxoid Diagnostic Reagents) unless significant differences in colony morphology and/or haemolysis intensity was observed, in which case additional colonies were also selected.

The current study received ethics approval from the Human Research Ethics Committee of the Northern Territory Department of Health and Menzies School of Health Research (approval 2015-2516).

### Whole genome sequencing and typing

Lancefield group C/G streptococcal isolates were retrieved from stored glycerol stocks kept at −70°C. Microbial DNA was extracted and 150bp paired-end libraries were prepared using the Illumina TruSeq prep kit. Sequencing was performed using the Illumina HiSeq X Ten platform (The Wellcome Trust Sanger Institute, United Kingdom). Fifty-four SDSE sequences were previously published by Xie et al.^9^ *S. pyogenes* sequences were previously described by Lacey et al.^13^ and available under Bioproject PRJNA879913.

Reads from Lancefield group C/G streptococcal isolates were checked for contamination using Kraken2 v2.1.2.^23^ Any sequences with >5% reads assigned to a species other than SDSE, with the exception of *S. pyogenes*, was excluded. Genomes were assembled using a previously described pipeline^9^.

*In silico* typing of the hypervariable N-terminal domain of the *emm* gene was performed using emmtyper v0.2.0 (https://github.com/MDU-PHL/emmtyper) and MLST assigned using MLSTv2.22.0 (https://github.com/tseemann/mlst)^24^. Genomic sequence clusters, representative of global SDSE populations, were assigned using PopPUNK v.2.60 with a scheme available at https://www.bacpop.org/poppunk/ (v1)^9, 25^. Antimicrobial resistance and virulence genes were inferred as previously described^9^. Genome metadata is available in Supplementary Table 1. *S. pyogenes* genomic sequences clusters were assigned with a scheme available at https://poppunk.net/pages/databases.html^26^.

The pangenomes for SDSE and *S. pyogenes* were constructed using Panaroo v1.2.10^27^ in ‘strict’ mode with initial clustering at 98% length and sequence identity followed by a family threshold of 70%. Core genes were defined as genes present in ϕ99% of genomes. Pangenome gene synteny was mapped using Corekaburra v0.0.5^28^.

Maximum likelihood phylogenetic trees for SDSE and *S. pyogenes* isolates were inferred using IQ-tree v2.0.6 with a GTR+F+G4 model and 1000 UFBoot replicates^29, 30^. Alignments for SDSE were generated using Snippy v4.6.0 (https://github.com/tseemann/snippy) against reference genome GGS_124 (NC_012891.1) and *S. pyogenes* against reference genome MGAS5005 (NC_007297.2) with MGE regions masked. Recombination was not masked. Maximum parsimony trees were inferred within genomic sequence clusters to validate predicted transmission clusters using phangorn v2.10.0 and SNP alignments generated by split kmer analysis (SKA v1.0)^31, 32^.

### MGE comparison

MGEs were systemically extracted from SDSE and *S. pyogenes* genomes using a pangenome synteny based approach based on proMGE and as previously described^9, 16^. The pipeline was modified to extract nucleotide sequences of accessory genomic segments classified as MGEs. Sequences from SDSE and *S. pyogenes* were initially clustered using CD-HIT^33^ v4.8.1 with a sequence identity threshold 0.8 and length difference cut-off 0.8. Clusters with sequences from both species were inspected and pairwise alignments generated using minimap2 v2.24^34^.

### Transmission clustering

Transmission clusters representing isolates predicted to have formed a recent transmission chain based on WGS data was determined using a SNP threshold of <8 and >99% shared gene content. A SNP threshold of <8 was determined using the maximal SNP distance between isolates of the same *emm* and MLST isolated within the same individual, a surrogate for SNP diversity within a single infecting strain. A gene content threshold of >99% determined from pangenome analysis was chosen to capture MGE gain/loss events.

Pairwise SNP distance between isolates within the same genomic sequence cluster was performed using SKA v1.0^31^ from reads using the ‘fastq’ command with default coverage cut-off 4, minimum minor allele frequency 0.2, minimum base quality 20 and kmer size 15. Single linkage clustering with a SNP distance of <8 was calculated using the ‘distance’ command. SKA requires exact kmer matches in order to detect SNPs between the flanking/split kmers and therefore may miss SNPs between more divergent sequences. The 150bp hypervariable N-terminal region of the *emm* gene poses such a challenge and has previously been shown to be able to undergo recombination. As such, isolates clustered by SKA were checked for matching *emm* types. In the case of different *emm* subtypes, alignments of the hypervariable region *emm* region were manually inspected to determine the number of SNPs. Finally, a pangenome comparison and single linkage clustering within each SKA cluster at 20 genes (approximates 99% gene similarity) was performed to generate the final transmission clusters.

Transmission cluster persistence was calculated by Kaplan-Meier estimation from time of first detection of a transmission cluster in a community to the first visit where the transmission cluster was not detected without subsequence re-appearance. The difference between SDSE and *S. pyogenes* was calculated by Cox Proportional Hazards as implemented in survival v3.4.0 and survminer v0.4.9.

### Household transmission network

Transmission networks between households in each community were inferred using a modified version of the model described by Lacey *et al.*^13^ and the R packages igraph v1.3.5 for network analysis, and ggraph v2.1.0, and scatterpie v0.1.8 for visualisation. Networks were initially inferred for SDSE and *S. pyogenes* separately. Transmission clusters predicted using WGS were mapped against epidemiological metadata to generate adjacency matrices from which transmission networks were inferred. Each household was represented by a node within the transmission network and unweighted edges (transmission events or links) were drawn between households, or within the same household, when unique individuals carried isolates from the same transmission cluster across successive community visits (transmission window between 12 to 44 days). Isolates could be linked to multiple other isolates within the same transmission cluster and respective transmission window. The isolate detected at the earlier community visit/time point was denoted as putative source for the purposes of predicting the contribution of throat and skin carriage/infection to transmission. Edges were drawn for each transmission window and assigned to the latter, ‘recipient’, community visit. As households were sampled in order over a short window (range 1–4 days) within each community visit, transmission edges were not inferred within the same community visit given the uncertainty in predicting a source.

To determine transmission overlap between SDSE and *S. pyogenes*, the intersection between the transmission networks for SDSE (Gsdse) and *S. pyogenes* (Gpyo) at each transmission window (w) was taken. Here, households were nodes and with undirected edges (E) drawn if they were linked by a transmission event. The percentage of shared edges (f) was calculated as:

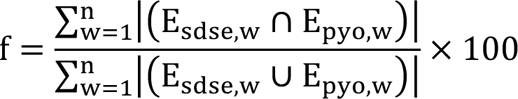

### Models of independent transmission

A null model of independent transmission was generated for the SDSE and *S. pyogenes* transmission networks by node-label permutation. The household labels of the transmission adjacency matrix generated from the observed SDSE data were permuted over 10,000 iterations and compared to the *S. pyogenes* transmission network inferred from observed data as described above. The number of overlapping edges at each transmission window was calculated, summed for each iteration, and compared to the observed number of shared transmission edges. A one-sided p-value testing the hypothesis of SDSE and *S. pyogenes* transmission interference was calculated by the proportion of permutations with shared edges ≤ observed shared edges.

A model of independent inter- and intra-species transmission for household co-occurrence was generated by permutation of positive SDSE and *S. pyogenes* swabs across individuals and households within each community visit while accounting for grouping of isolates from the same transmission cluster within households. Sequenced isolates from the same transmission cluster and household-visit (159/609 SDSE and *S. pyogenes* combined, 26%) were collapsed into a single positive result (159 collapsed to 69 positive swabs). Positive SDSE and *S. pyogenes* swabs at each community visit were then permuted across all individuals sampled at that respective community visit. A co-occurrence within a household was counted when SDSE and *S. pyogenes* were present simultaneously in individuals in a household regardless if they were from the same individual or across multiple individuals. After 10,000 iterations, a one-sided p-value testing the hypothesis of SDSE and *S. pyogenes* transmission interference at a household level was calculated by the proportion of permutations with co-occurrences ≤ observed co-occurrences.

## Data availability

Accessions for newly sequenced SDSE isolates are listed in Supplementary Table 1 under the BioProject identifier PRJEB35476.

## Code availability

Scripts used to generate the transmission networks and null models are available at https://github.com/OuliXie/SDSE_transmission. Scripts for MGE extraction and classification from the pangenome are updated from that described previously^9^ and are available at https://github.com/OuliXie/Strep_MGE_pipeline.

## Acknowledgements

We thank the participants, communities, councils, Aboriginal research officers and health centres for their involvement in the original surveillance study. We thank Ross M. Andrews for his role in the original surveillance study. We acknowledge the assistance of the sequencing and pathogen informatics core teams at the Wellcome Sanger Institute, UK where this work was supported by the Wellcome Trust core grants 206194 and 108413/A/15/D. OX was supported by the Australian Health and Medical Research Council (NHMRC) postgraduate scholarship (GNT2013831) and Avant Foundation Doctors in Training Research Scholarship (2021/000017). MRD was supported by a University of Melbourne CR Roper Fellowship.

## Contributions

OX worked on study design, analysis, data interpretation and manuscript preparation. MRD and SYCT contributed to conception of the project and data interpretation. CZ, GTH, DJP and JAL contributed to transmission model design and data interpretation. JMM, MIM, ACB, PMG, BJC, JRC, and DCH contributed to data collection and curation. SDB contributed to genomic sequencing. All authors contributed to manuscript preparation and review.

