## Supplementary Figure 1-9 for "Overlapping transmission of group A and C/G *Streptococcus* facilitates inter-species mobile genetic element exchange"

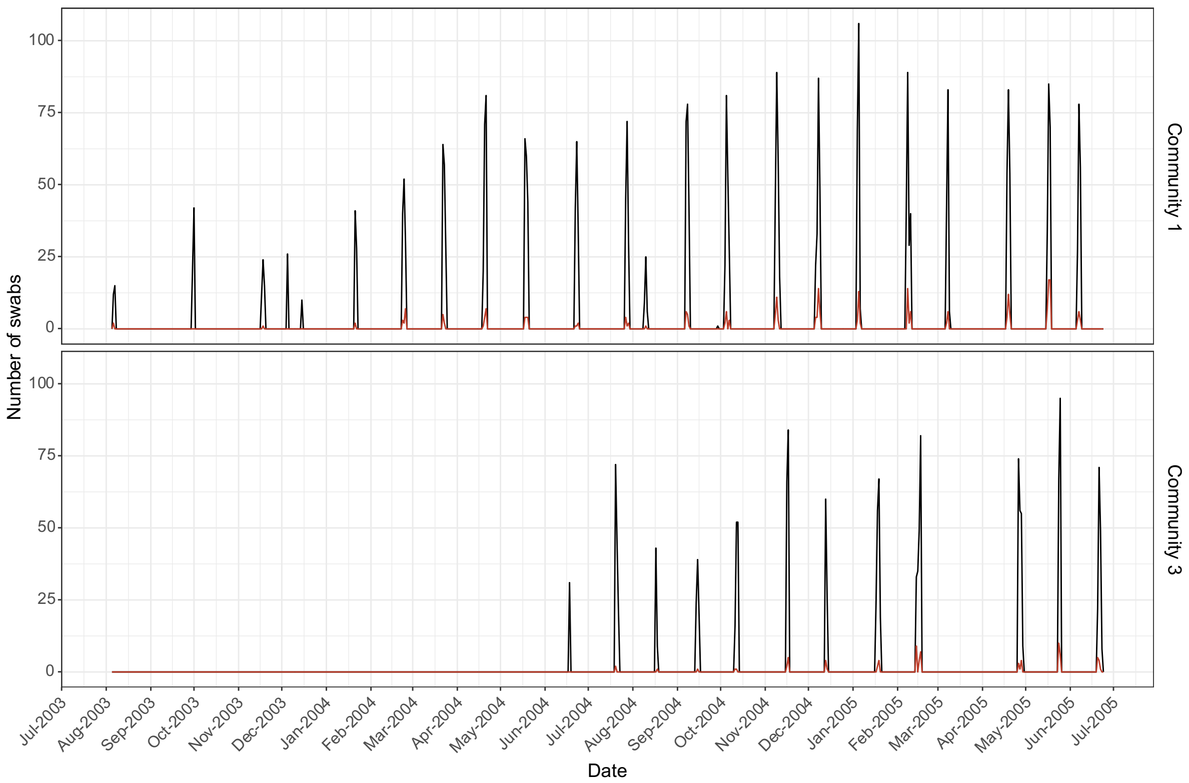


**Supplementary Figure 1.** Frequency and number of swabs during each community. The total number of swabs are represented by the black line and swabs positive for *Streptococcus dysgalactiae* subsp. *equisimilis* are represented by the red line. Communities were visited on approximately monthly intervals except during periods of extreme weather or community cultural events. Community 3 commenced recruitment in June 2004 after a previous community was replaced in the study due to low recruitment.

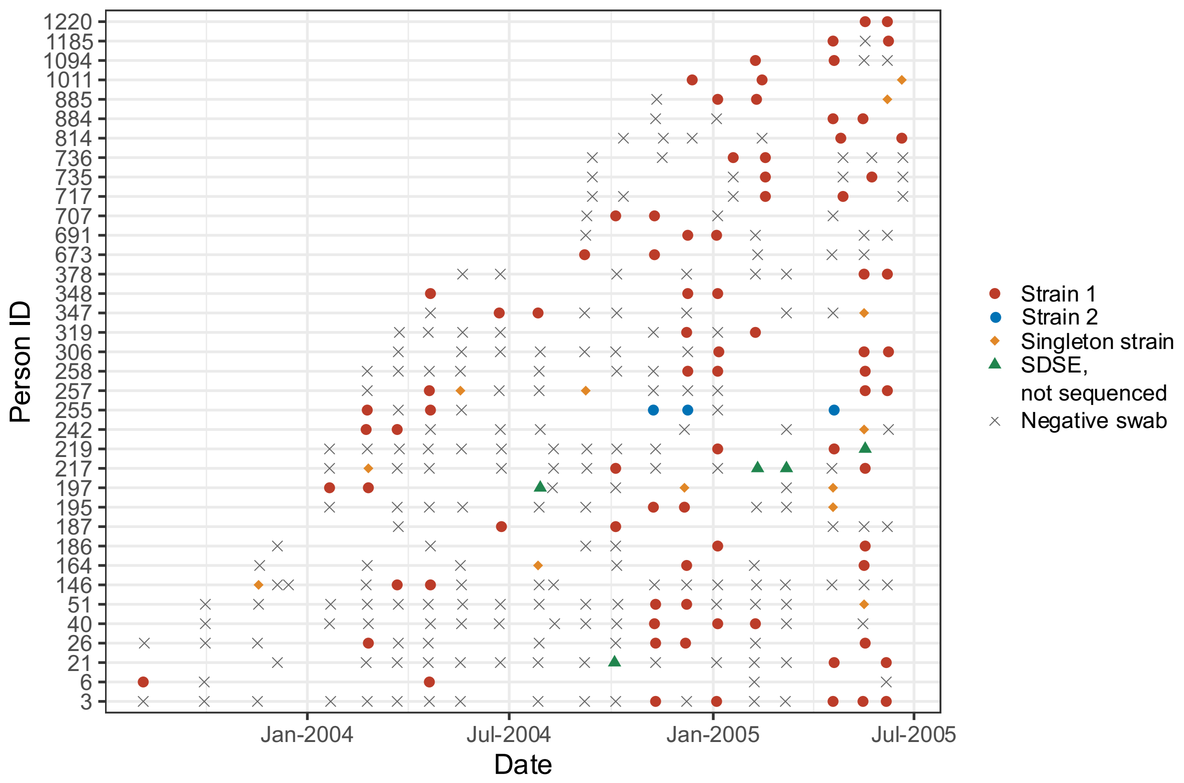


**Supplementary Figure 2.** Timeline of 36 individuals with SDSE isolated on multiple occasions. The same strain as defined by *emm* type and MLST within each individual are indicated by red or blue points. Strains which only appear once in each individual are indicated by yellow diamonds. Crosses denote occasions where a swab was taken but culture was negative. One individual had isolation of one isolate on three occasions and a different isolate on two occasions. Isolates were generally found on consecutive visits indicative of persistent carriage or separated by 1-2 negative swabs except for three individuals where the same isolate was found more than one year apart suggestive of reinfection.

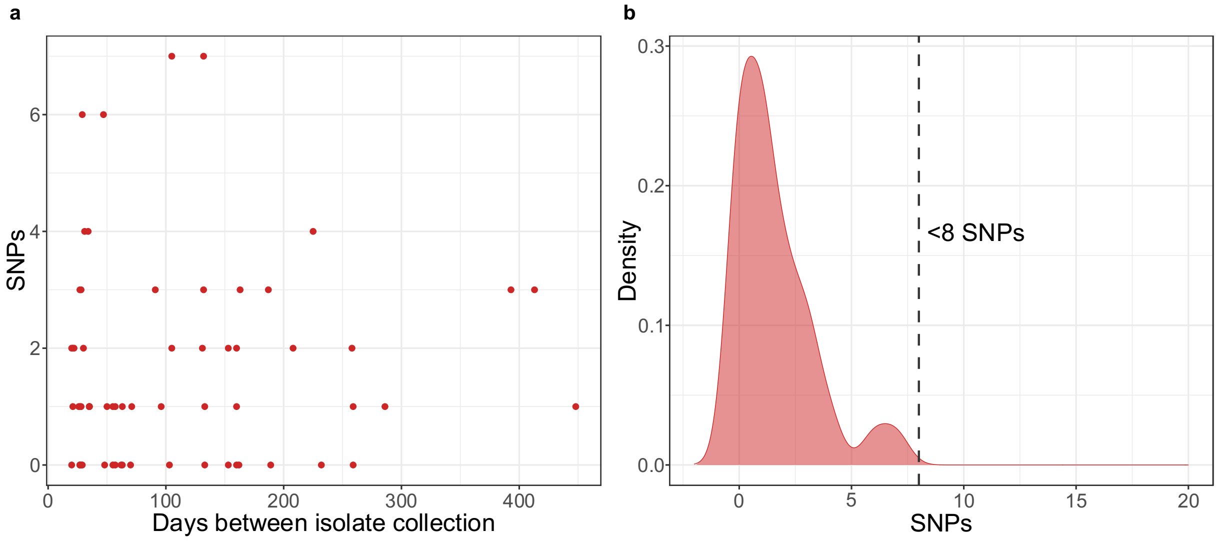


**Supplementary Figure 3.** Single nucleotide polymorphism (SNP) distance between isolates of the same *emm* and multilocus sequence type isolated from the same individuals. **a)** The SNP distance of 64 pairwise comparisons between 49 isolates. **b)** Density plot of SNP distances demonstrating a maximal SNP distance of <8 SNPs.

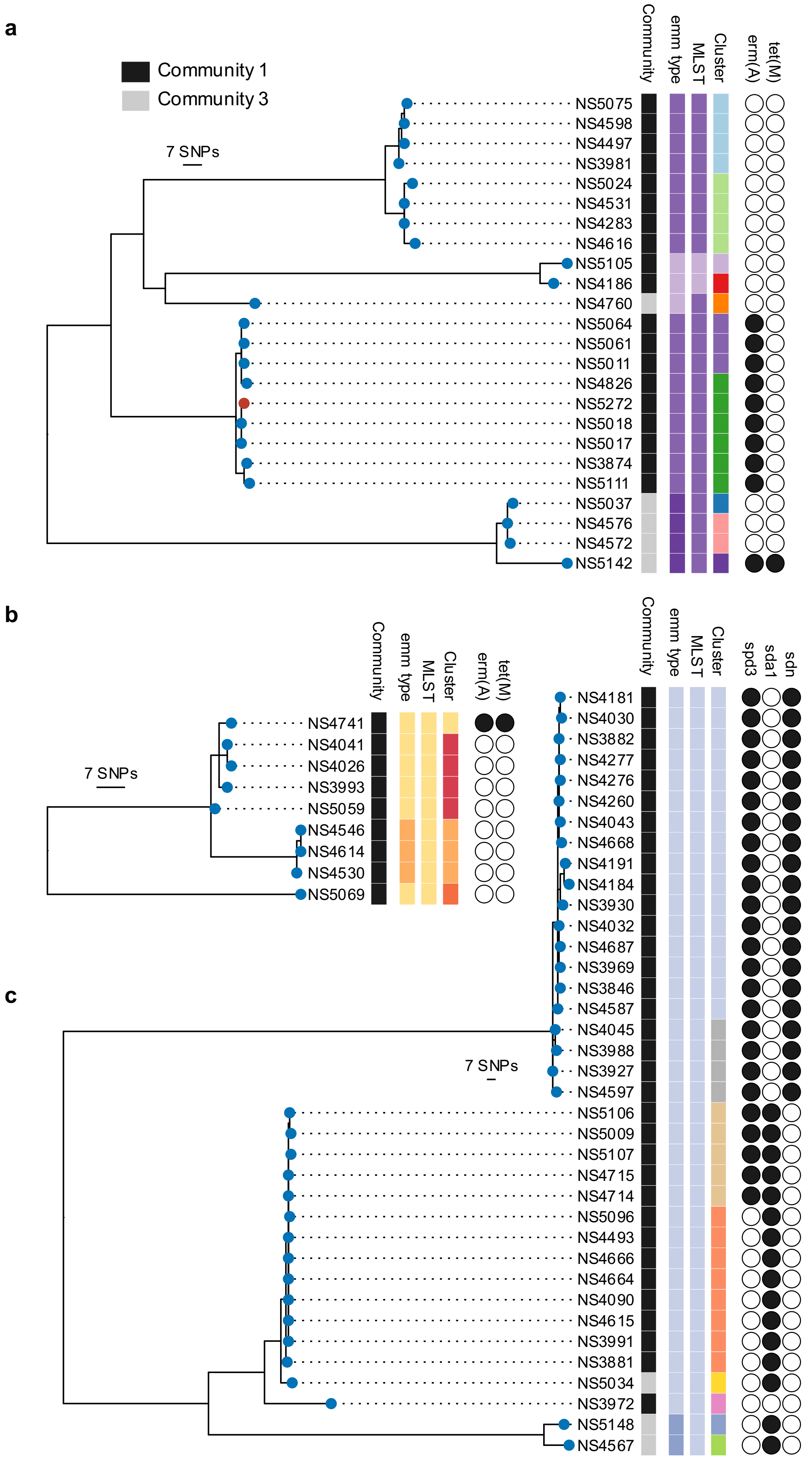
a
Community 1
Community 3
7 SNPs
Community
emm type
MLST
Cluster
erm(A)
tet(M)
NS5075
NS4598
NS4497
NS3981
NS5024
NS4531
NS4283
NS4616
NS5105
NS4186
NS4760
NS5064
NS5061
NS5011
NS4826
NS5272
NS5018
NS5017
NS3874
NS5111
NS5037
NS4576
NS4572
NS5142
b
7 SNPs
Community
emm type
MLST
Cluster
erm(A)
tet(M)
NS4741
NS4041
NS4026
NS3993
NS5059
NS4546
NS4614
NS4530
NS5069
c
7 SNPs
Community
emm type
MLST
Cluster
spd3
sda1
sdn
NS4181
NS4030
NS3882
NS4277
NS4276
NS4260
NS4043
NS4668
NS4191
NS4184
NS3930
NS4032
NS4687
NS3969
NS3846
NS4587
NS4045
NS3988
NS3927
NS4597
NS5106
NS5009
NS5107
NS4715
NS4714
NS5096
NS4493
NS4666
NS4664
NS4090
NS4615
NS3991
NS3881
NS5034
NS3972
NS5148
NS4567

**Supplementary Figure 4.** Maximum parsimony phylogenies of select *Streptococcus dysgalactiae* subspecies *equisimilis* (SDSE) genomic sequence clusters using core single nucleotide polymorphism (SNP) alignments. **a)** Genomic sequence cluster 2 contained three distinct *emm* subtypes *stC*6979.0 (3 light purple isolates), *stC*74A.0 (17 purple isolates), and *stG*2078.0 (4 dark purple isolates) from two multilocus sequence types (MLSTs) 17 (22 purple isolates) and 137 (2 light purple isolates). Isolates were separated into 10 distinct transmission clusters (including 5 singletons) which were mutually exclusive across the two communities. Isolates exhibited variable presence of macrolide resistance gene *erm(A)* and tetracycline resistance gene *tet(M)* or *erm(A)* alone on two distinct composite integrative conjugative element (ICE)-like accessory genomic segments. **b)** Genomic sequence cluster 30 contained two *emm* subtypes *stC*6979.0 (6 yellow isolates) and *stG*480.8 (3 orange isolates) from the same MLST 140. Isolate NS4741 was distinct from other closely related strains due to acquisition of a ~60kb composite ICE-like accessory genomic segment carrying the macrolide resistance *erm(A)* and tetracycline resistance *tet(M)* genes. The composite ICE-like segment was inserted in the same genomic region and shared 97% nucleotide identity to the *erm(A)*/*tet(M)* element from NS5142 in genomic sequence cluster 2. **c)** Genomic sequence cluster 9 contained two *emm* subtypes *stC*36.0 (35 light blue isolates) and *stG*6792.0 (two dark blue isolates), all from MLST 4. *stG*6792.0 isolates were separated into two distinct populations approximately 500 SNPs distant and *stC*36.0 isolates were approximately the same distance from the closest *stG*6792.0 population. Otherwise closely related isolates were separated by variable presence of prophage elements encoding *spd3*, *sda1*, and/or *sdn*.

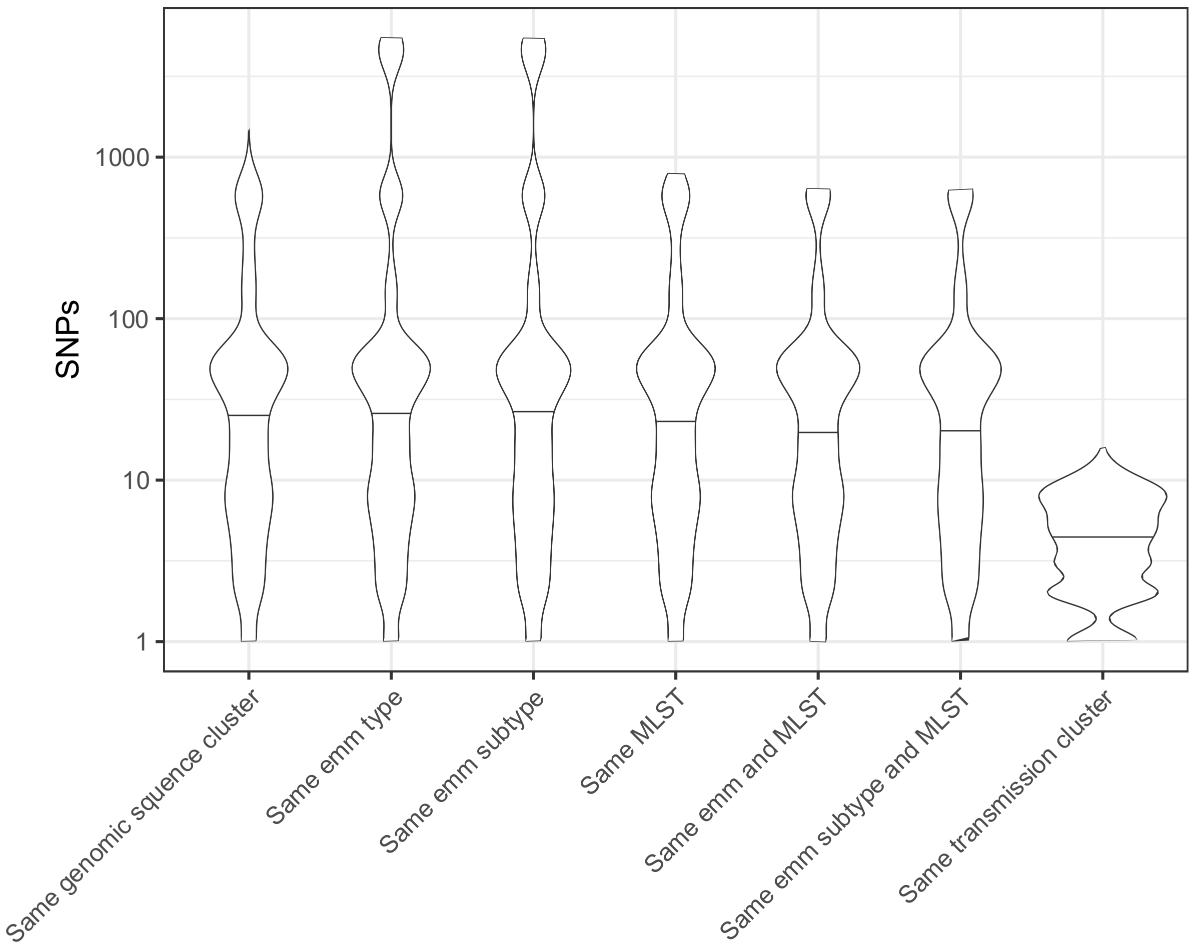


**Supplementary Figure 5.** Violin plot of *Streptococcus dysgalactiae* subspecies *equisimilis* (SDSE) pairwise single nucleotide polymorphism (SNP) distances on a $\log_{10}$ scale between isolates across different categories of classification. Genomic sequence clusters were determined by PopPUNK[25] as described previously using a global SDSE database[9]. The distribution of SNP distances in transmission clusters using single linkage clustering at threshold of <8 SNPs and >99% shared gene content provided the greatest resolution in determining recent transmission clusters. The presence of the same *emm* or *emm* subtype in isolate pairs with >5000 SNPs distance was indicative of carriage of the same *emm* and *emm* subtype across very different genomic backgrounds/lineages.

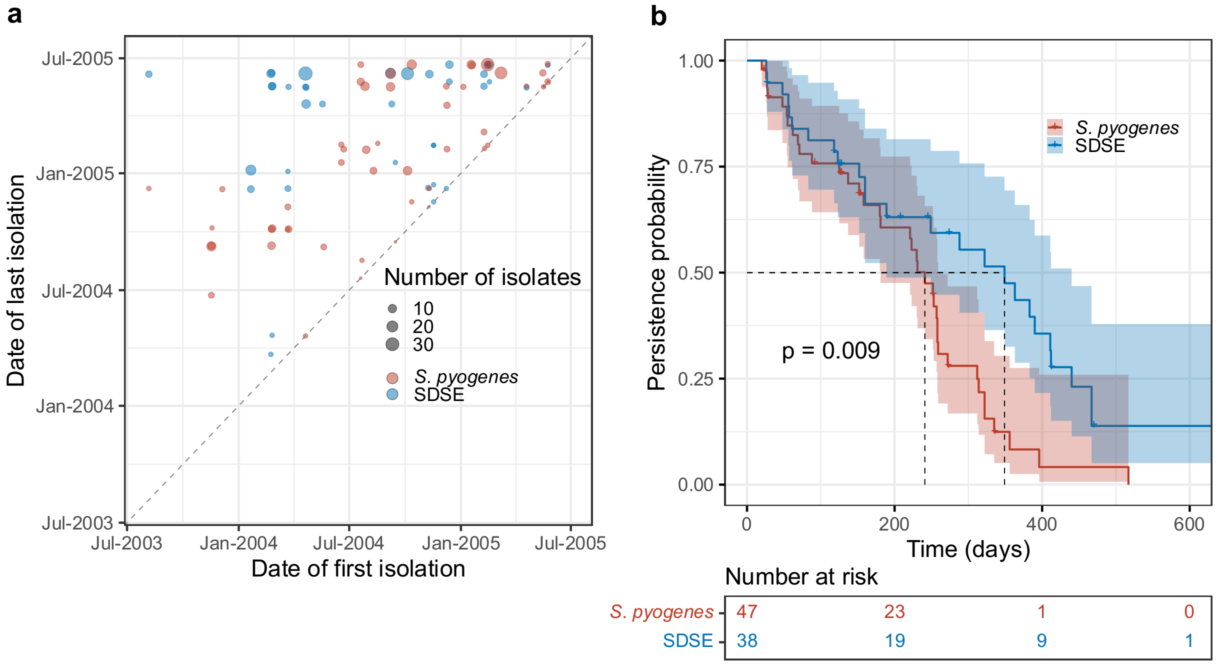


**Supplementary Figure 6.** Transmission cluster persistence of *Streptococcus dysgalactiae* subspecies *equisimilis* (SDSE) compared to *Streptococcus pyogenes*. **a)** Date of last detection of a transmission cluster against date of first isolation. Dotted identity line represents isolates which were detected at only a single time point. The further from the identity line, the longer a transmission cluster persisted while those present at the top of the chart were present at the last study visit in July 2005. **b)** Survival curve of transmission cluster persistence. SDSE transmission clusters persisted for a median of 349 days (95% CI 189-440 days) while *S. pyogenes* transmission clusters persisted for a median of 241 days (95% CI 181-259 days). SDSE transmission clusters persisted significantly longer than *S. pyogenes* (log-rank p = 0.009).

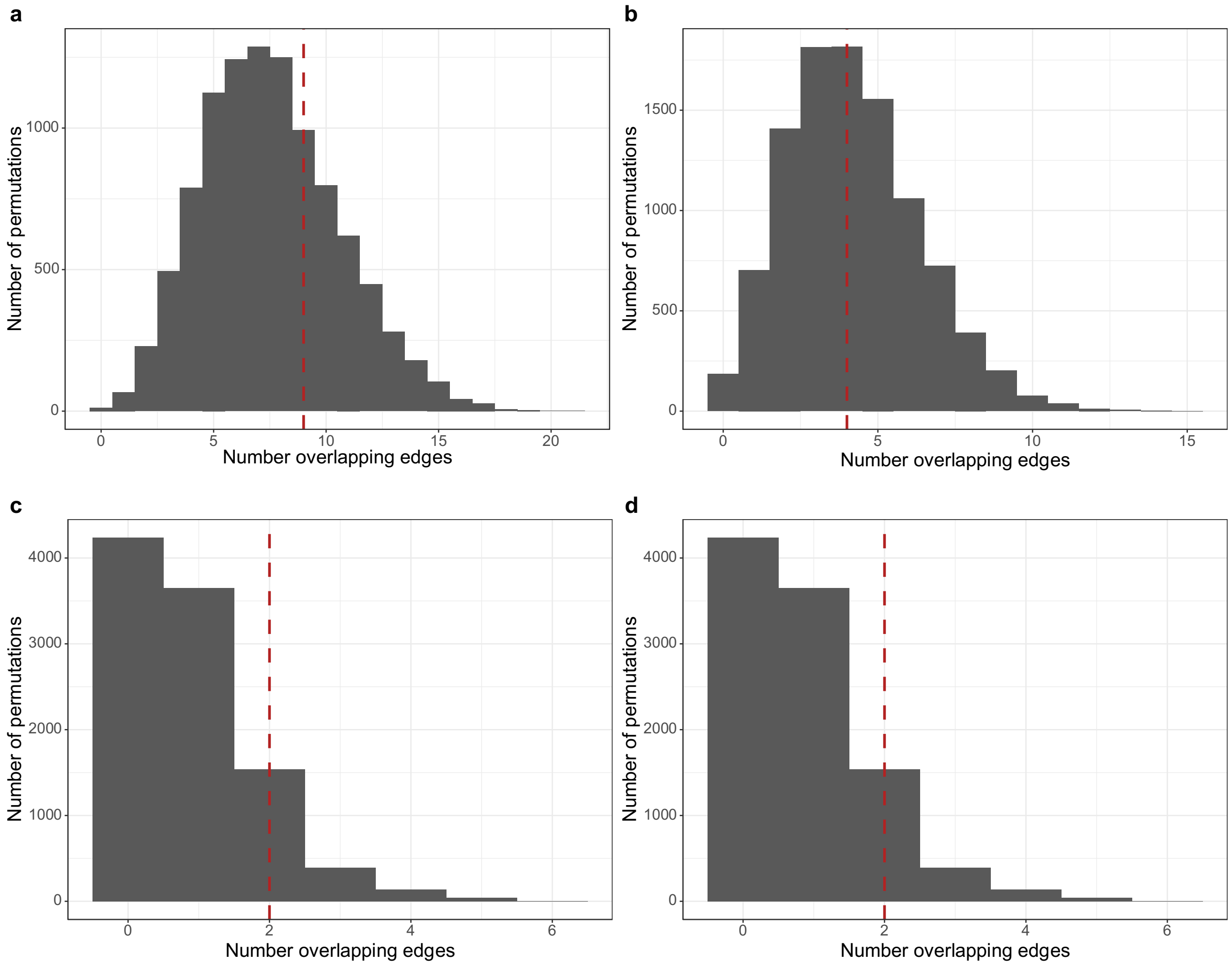


**Supplementary Figure 7.** Distribution of undirected shared transmission edges between *S. dysgalactiae* subsp. *equisimilis* (SDSE) and *S. pyogenes* in models of independent transmission. Null models were generated by node label permutation of transmission network matrices and repeated for 10,000 iterations. The observed number of overlapping edges are marked by red dashed lines. **a)** Community 1 one-sided p-value ≤ observed value = 0.75. **b)** Sensitivity analysis of community 1 limited to SDSE and *S. pyogenes* throat swabs only. One-sided p-value ≤ observed value = 0.59. **c)** Community 3 one-sided p-value ≤ observed value = 0.94. **d)** Sensitivity analysis of community 3 limited to SDSE and *S. pyogenes* throat swabs only. One-sided p-value ≤ observed value = 0.94.

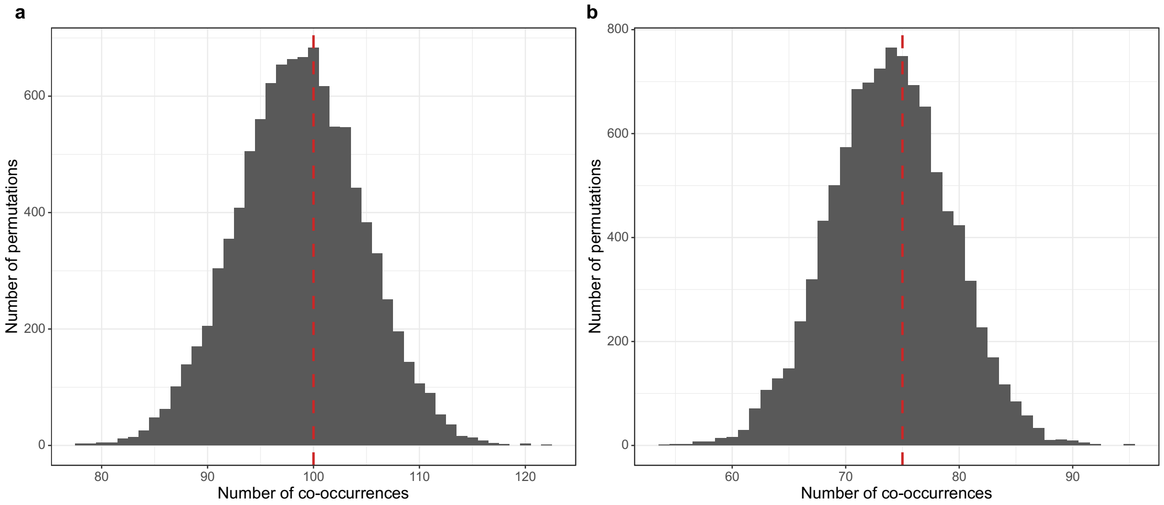


**Supplementary Figure 8.** Distribution of household level co-occurrences of *S. dysgalactiae* subsp. *equisimilis* (SDSE) and *S. pyogenes* across both communities in models of independent transmission. Null models were generated by permutation of swabs across sampled individuals within the same community visit after correcting for clustering of isolates from the same transmission cluster within households. Permutations were repeated 10,000 times. Observed frequency of co-occurrence of the two species at household-visits are denoted by red dashed lines. **a)** Inclusion of both skin and throat isolates generated a one-sided p-value ≤ observed value = 0.62. **b)** Sensitivity analysis with only isolates from throat swabs generated a one-sided p-value ≤ observed value = 0.62.

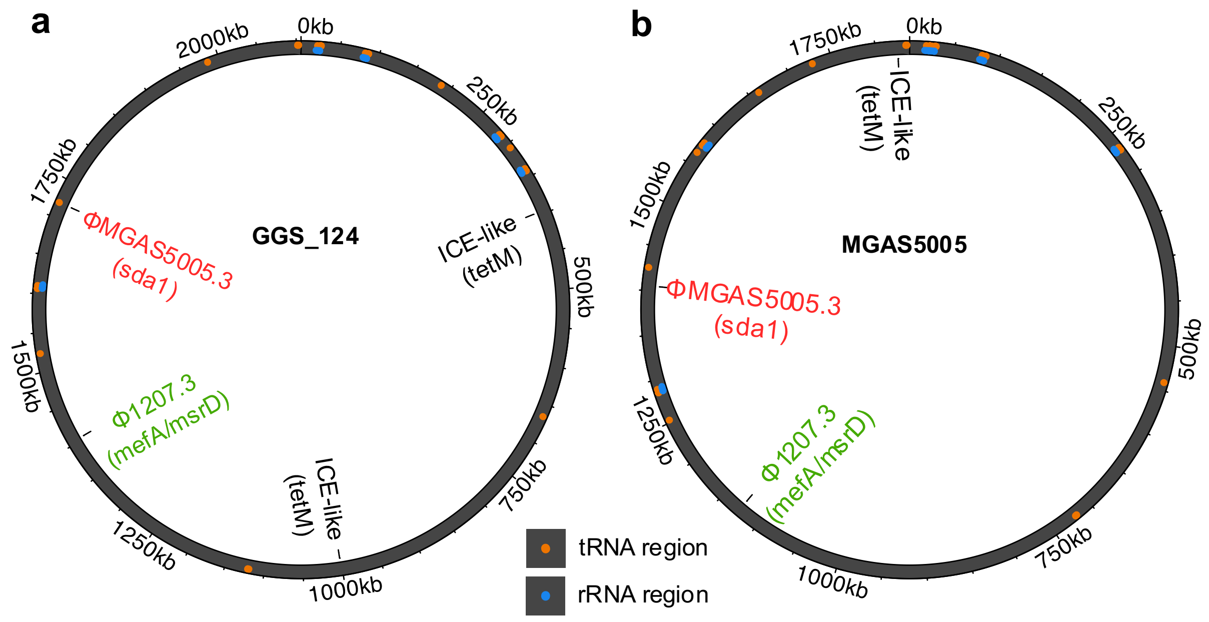


**Supplementary Figure 9.** Genome representations of **a)** *Streptococcus dysgalactiae* subsp. *equisimilis* (SDSE) reference GGS_124 NC_012891.1 and **b)** *Streptococcus pyogenes* reference MGAS5005 NC_007297.2 with mapped insertion of regions of three cross-species shared mobile genetic elements (MGEs) found in this dataset of co-circulating isolates. Prophage ϕ1207.3 carrying the macrolide resistance genes *mef(A)*/*msr(D)* was present at a cross-species conserved insertion region (mapped to SDEG_RS05910-SDEG_RS05915 in GGS_124 and M5005_RS05690-M5005_RS05690 in MGAS5005). Prophage ϕMGAS5005.3 was also present at a cross-species conserved insertion region (mapped to SDEG_RS08710-SDEG_RS08720 in GGS_124 and M5005_RS07010-M5005_RS0720 in MGAS5005) as described previously[9]. An ICE-like element carrying *tet(M)* was present at two different insertion regions in SDSE (SDEG_RS02085-SDEG_RS02090 and SDEG_RS05020-SDEG_RS05030) and a separate insertion region in *S. pyogenes* (M5005_RS09125-M5005_RS09130).

**Supplementary Table 1. a)** List of 294 *Streptococcus dysgalactiae* subspecies *equisimilis* (SDSE) isolates with whole genome sequencing data included in the study with associated clinical and genomic metadata. **b)** Virulence and antimicrobial resistance gene presence and absence across the 294 SDSE isolates.

*See separate excel file*
